# Using the English national health service dataset for research into mental health service use among children and young people

**DOI:** 10.1101/2025.11.03.25339389

**Authors:** Niloofar Shoari, Kate Lewis, Ruth Blackburn, Benjamin Ritchie, Steven Cummins, Nina T Rogers, Samantha Hajna, Pia Hardelid

## Abstract

**Background:** Routinely collected administrative data, such as the Mental Health Services Data Set (MHSDS) in England, provide opportunities to investigate determinant and patterns of mental health service use across the population. Persistent challenges with data completeness and consistency, however, limit their value for robust population-level research.

**Objective:** To assess the methodological robustness and research utility of the MHSDS for investigating patterns and determinants of mental health service use among children and young people (CYP) in England between 2016 and 2023.

**Methods:** We evaluated the completeness and consistency of key sociodemographic variables (gender, ethnicity, location, and socioeconomic status) over time and across local authorities. To assess the impact of data quality on analytical validity, we modelled the likelihood of care contact attendance using sociodemographic covariates, comparing estimates from complete-case analyses with those from models treating missing or conflicting data as distinct categories. Spatial and temporal patterns of CYP in contact with (referred to) mental health services and attendance rates were examined annually and by local authority.

**Findings:** From 2016 to 2023, over 4.7 million CYP were in contact with mental health services, of whom 62.4% had at least one attended contact. Missing data were substantial, persistent, often co-occurred in subpopulations with distinct attendance patterns. The proportion of CYP referred to services increased from 1.9% (371,655) in 2016 to 9.7% (1,699,899) in 2023, while attendance rates remained stable at 59.8%. Spatial analyses highlighted regional variations, with higher attendance rates in Northern England

**Conclusions:** Data completeness influences the validity of research using the MHSDS. Addressing this issue is essential for producing equitable and reproducible evidence in CYP mental health research.

**Clinical Implication:** Methodological improvements in handling missing data and temporal inconsistencies will strengthen the interpretability, reproducibility, and equity relevance of MHSDS-based mental health research.

## Introduction

The prevalence of mental health conditions among children and young people (CYP) has risen steadily worldwide ^1,2^. In the United Kingdom, the Survey of Mental Health of Children and Young People reported an increase in self-reported probable mental disorders among CYP, with prevalence rates approximately doubling from 11.5% in 2017 to 22.7% in 2023 ^3^. This upward trend aligns with global data, particularly after the COVID- 19 pandemic ^4–6^. The growing prevalence highlights the need for understanding how mental health services are utilised to meet the increasing demand among CYP.

Despite this need, evidence on how CYP utilise mental health services remains limited. Mental health service utilisation refers to any point of contact that patients have (receiving or awaiting treatment) with in and out-patient mental health services. Barriers to receiving care, such as high rates of referral rejection, long waiting lists, and social stigma, and material disadvantage can delay care and contribute to unequal access to treatment, even for those CYP who are referred to services ^7^. Addressing how these barriers manifest across sociodemographic groups is critical for improving service provision and policy planning.

There is a lack of comprehensive knowledge about the spatial and temporal patterns of service utilisation, and how these vary across different sociodemographic groups, including gender, ethnicity, and socioeconomic status. Such information is crucial for designing equitable and effective mental health care systems and making informed policy decisions. However, the reliability of such analyses is inherently dependent on the quality and completeness of the underlying data.

The Mental Health Services Dataset (MHSDS), commissioned by the English National Health Service (NHS), represents a valuable national resource that is originally collected to monitor and support the delivery of mental health, learning disabilities, and autism services Its primary purpose is to facilitate service planning and improve the quality of care by capturing detailed information about individuals in contact with these services.

Although not originally intended for research, the MHSDS provides a rich source of individual-level data, capturing detailed information on various aspects of care, including sociodemographic characteristics (e.g., age, gender, ethnicity), referral details (e.g., referral source, reason, team referred to), clinical data (e.g., diagnoses and care plans), and patterns of service utilisation (e.g., duration of care and type of service provided). This wealth of information makes the MHSDS invaluable for understanding who accesses mental health services, identifying barriers to treatment, and examining how these factors vary across time and geography. Furthermore, the dataset enables researchers to explore service uptake across different regions and populations, highlighting service gaps and inequalities in access. It also supports evaluation of the effectiveness of specific policies and interventions on mental health service use, contributing to evidence-based policy and practice.

Several studies have used the MHSDS to examine various aspects of mental health care. For example, a cross-sectional analysis found that females, younger CYP, and individuals with mixed ethnicity, or specific conditions such as substance use, eating disorders, self-care difficulties had a higher number of care contacts (compared to an average of 11.12 care contacts), while another study found that CYP with more severe problems had shorter waiting times for mental health services ^8^ ^9^. A longitudinal analysis focusing on pregnant women showed a decrease in acute care referrals and an increase in community care use following the introduction of Community Perinatal Mental Health Teams (CPMHTs) ^10^. These examples illustrate the dataset’s potential for informing mental health research and service planning.

Despite its utility, the MHSDS also presents challenges as a research dataset. As it was not originally collected for research purposes, the MHSDS requires extensive processing to address issues such as data quality, harmonisation, and integration across multiple separate data tables ^11^ ^12^. Local variation in service provides and delivery further influences patterns of service use, but the MHSDS does not capture the underlying service models driving these differences, potentially affecting the interpretation of findings. Addressing these limitations is critical to ensuring robust, reliable research outcomes.

Our study is an observational analysis of routinely collected administrative data that examines the methodological robustness and research utility of the MHSDS for investigating CYP mental health service use in England. We aim to:

- Assess the completeness and consistency of key sociodemographic variables including ethnicity, gender, location, socioeconomic status, and determine whether data quality issues vary systematically across sociodemographic groups.
- Evaluate how data quality influences analytical validity and reliability of research findings.
- Characterise spatial and temporal variations in mental health service utilisation across English local authorities and over time, stratified by sociodemographic factors, including age, ethnicity, gender, socioeconomic status.

## Methods

### Data source, study population and period

The MHSDS (version 6), held and maintained by NHS England, consists of 61 tables compiling data from NHS-funded mental health service providers. These tables provide information on patient demographics, referrals, care activities, group sessions, Mental Health Act (MHA) episodes, hospital spells, and clinically coded diagnoses (using the International Classification of Diseases version-10 ICD-10), staff roles (e.g., primary consultant specialty), service settings (e.g., inpatient, outpatient, community care). The Mental Health Act (MHA) refers to the legislation governing the assessment, treatment, and rights of individuals with mental health conditions who require inpatient care, sometimes under compulsory admission. Data are submitted by individual providers to a central NHS registry, ensuring a standardized national dataset for monitoring and planning mental health services.

MHSDS v6.0 follows a monthly submission process, based on the financial year (April to March). Each activity period records all NHS-funded services provided within the month, such as care contacts (e.g., outpatient appointments, therapy sessions, community visits). Providers are required to submit data in a given month by the end of the following month. The multiple submission window model allows updates or corrections to prior submissions within the same financial year, ensuring errors or missed submissions can be addressed ^13^.

Our analysis was restricted to CYP residing in England aged 0-25 years inclusive at the start of each financial year.

### Outcomes

Our outcomes of interest included

- CYP who were referred to mental health services from any source or admitted to inpatient psychiatric hospitals (referred to as CYP in contact with mental health services herein)
- CYP in contact with mental health services who attended at least one care contact (referred to as appointment attendance herein)
- CYP who were referred to mental health service with no recorded or attended care contact

These were calculated for each English local authority (LA) and financial year, using the 2011 boundary definition, with 327 LAs included. Local authorities are responsible for public services such as schools, social care, and monitoring public health, making them a key geography for analysing mental health service use for CYP.

The proportion of CYP in contact with services was calculated as the number of CYP in contact with services divided by the total population of the same age group within each LA for that year. The latter was obtained from the Office for National Statistics (ONS)^14^. The proportion of CYP attending care appointments was calculated as the number of CYP who attended at least one care contact divided by those who were in contact with services. The third outcome includes CYP without linked care contacts who are those with rejected referrals, missed appointments, or those still waiting for a decision on their referral. See supplemental file for details of data structure and integration.

### Covariates

Covariates include age, ethnicity, gender, healthcare provider, Lower Super Output Areas (LSOA), local authority, and neighbourhood level socioeconomic status defined by the Index of Multiple Deprivation (IMD). Socioeconomic status, LSOA, and local authority were derived from the patient’s postcode of residence. LSOAs are geographical units in England used to report Census small area statistics. Each LSOA contains between 400 and 1,200 household or a population of 1,000 to 3,000 people ^15^.

Age was categorized as 0-4, 5-9, 10-14, 15-19, and 20-25 years. Ethnicity was categorized using high-level classifications, including White, Black or Black British, Asian or Asian British, Mixed, and Other Ethnic Groups, with inconsistent or missing data appropriately flagged. Socioeconomic status and residential locations were derived from patient postcodes, enabling analyses based on local deprivation levels. The healthcare provider refers to the organisation or practitioner delivering care and was identified through the sociodemographic table of MHSDS. Retrieving consistent information on residential locations in MHSDS was not straightforward due to address changes between records. When an individual had multiple LSOA/local authority codes registered in each financial year, we counted the subject in all relevant LSOAs/local authorities. The IMD, recorded as quartiles (from most to least deprived areas), has been available in the MHSDS since 2019. Details on covariates are available in the supplemental file.

### Statistical Analyses

Figure 1 summarises the MHSDS tables used in this study, outcomes and analytical framework.

**Figure 1.**
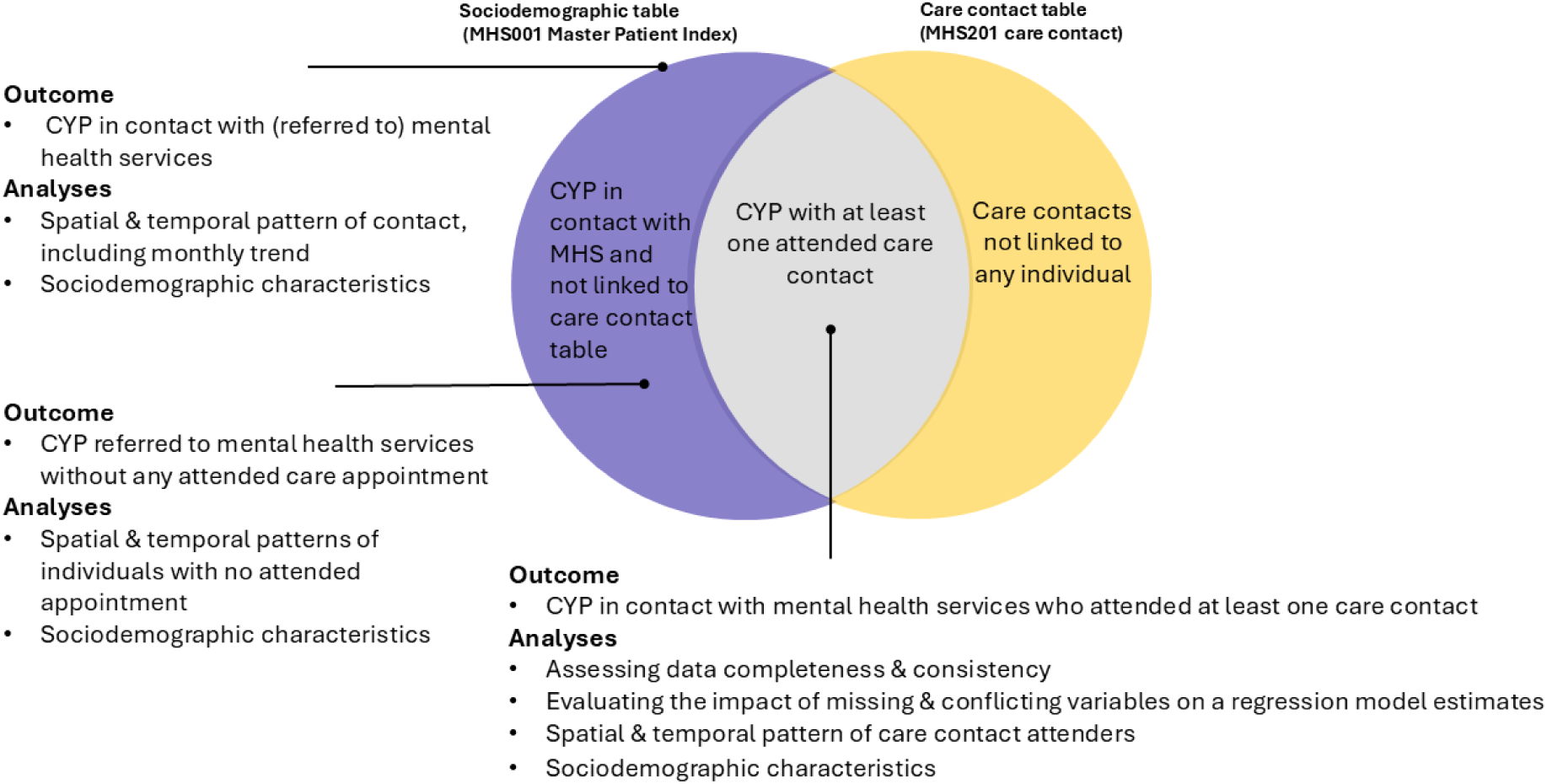
Diagram of the outcomes under study and analytical methods applied.

#### Assessing data completeness and consistency

We examined the proportion of CYP with missing or conflicting information on gender, ethnicity, neighbourhood-level socioeconomic status, and LSOA of residence, tracking changes in data quality over financial years. Since healthcare providers did not all begin submitting data at the same time, and the number of providers varied annually, we considered the number of providers each year. To identify structural sources of missingness, we examined variations by local authority and health care provider. We also explored dependencies between variables to determine whether data gaps were systematic. For example, we looked at age and ethnicity distributions for missing gender, and gender and age for missing ethnicity. For missing LSOA data, we assessed the proportion of missing records across gender, ethnicity, and age groups.

#### Evaluating the impact of missing and conflicting data

To assess how data quality affects analytical validity, we modelled the number of attended care contacts as a proportion of the total care contacts for each CYP using a binomial regression with age, ethnicity, gender, and neighbourhood-level socioeconomic status as covariates. Details on data preparation for the model are explained in the supplemental file.

Analyses were conducted for 2023 using two complementary approaches: (i) missing and conflicting gender, ethnicity, and IMD are considered as distinct categories, (ii) complete-case analysis, where CYP with missing or conflicting information are excluded. Differences in coefficient estimates and 95% confidence intervals were compared to evaluate bias introduced by missing and conflicting information. Bootstrap resampling (1,000 iterations) was used to examine coefficient stability and quantify uncertainty attributable to incomplete data.

#### Spatial and temporal patterns of service use

We examined the spatial and temporal patterns of CYP in contact with mental health services and care contact attendance, stratified by financial year (2016–2023) and local authority. Monthly submission data were further examined to assess the influence of reporting cycles and dataset updates on observed patterns and data reliability.

#### Sociodemographic profiling

This analysis explored sociodemographic characteristics (age, gender, ethnicity, IMD) associated with three outcomes: CYP in contact with service, those with at least one attended care contact, and those without attended contacts. This investigation provided context for interpreting utilisation patterns and highlighted potential inequities in engagement.

#### Sensitivity analysis

We conducted sensitivity analysis to assess the consistency and robustness of our findings. Specifically, we repeated the spatial and temporal pattern analysis using data exclusively from healthcare providers who consistently submitted their records from 2016 to 2023 to NHS digital. Healthcare providers were included if they submitted data for each year of the study period, regardless of record volume. We also performed the binomial regression analysis using data from 2021 year to assess the consistency of the results across different years.

## Results

Between 2016 and 2023, over 4.7 million CYP in England were in contact with mental health services. Of these, 62.4% (2.9 million) attended at least one care contact.

### Data completeness and consistency

Data completeness was strongly influenced by when and how health providers submitted their records over time. The number of reporting mental health providers increased from 96 in 2016 to 416 in 2023, affecting absolute counts but not spatial and temporal trends. In 2023, 23.9% (n=406,084) of CYP in contact with mental health services lacked ethnicity data, 4.4% (n=74,539) missing gender, and 11.4% (n=194,868) missing neighbourhood socioeconomic status (IMD). Among CYP with at least one attended care contact in 2023, missingness was similar (4.8% gender, 18.7% ethnicity, and 13.3% IMD). Detailed summaries of missing and conflicting records for CYP in contacts with services and those with at least one attended care contact over the study period (2016–2023) are reported in Supplemental Table 1 and Table 2, respectively.

Missingness pattern was not completely at random. Across years, CYP with missing gender data were often missing ethnicity, geographical information, and were disproportionately from the most deprived neighbourhoods (i.e., 1^st^ quartile of IMD). For example, in 2023 about half of those with missing gender (n=11,520) also lacked ethnicity data, and 31.2% (n=7,225) lived in the most deprived neighbourhoods. CYP with missing ethnicity were predominantly females from the most deprived neighbourhoods. Missing LSOA data ranged from 3.2% (n=5,454) in 2017 to 27.64% (n=95,903) in 2020, largely from independent and non-NHS providers. IMD data were unavailable before 2019 and inconsistently reported thereafter. Conflicting gender or ethnicity entries were rare (<1%) and excluded from spatial analyses. Characteristics of CYP with missing information are detailed in Supplemental Table 3.

### Impact of missing and conflicting data on model estimates

Regression models showed that missing ethnicity and gender data were associated with attendance patterns in opposite directions compared with other categories. CYP with missing gender and ethnicity had lower odds of attending care contacts (OR=0.57, 95%CI: 0.56,0.59, and OR= 0.67, 95%CI:0.66, 0.68, respectively). In contrast, missing information on neighbourhood deprivation (IMD) was associated with higher odds of attendance (OR=1.37, 95%CI:1.35,1.38) (Figure 2). The magnitude and direction of coefficients were broadly consistent between the complete-case analysis and the model treating “missing” and “conflicting” data as explicit categories (supplemental Figure 1). Bootstrap analysis further confirmed the stability of the findings.

**Figure 2.**
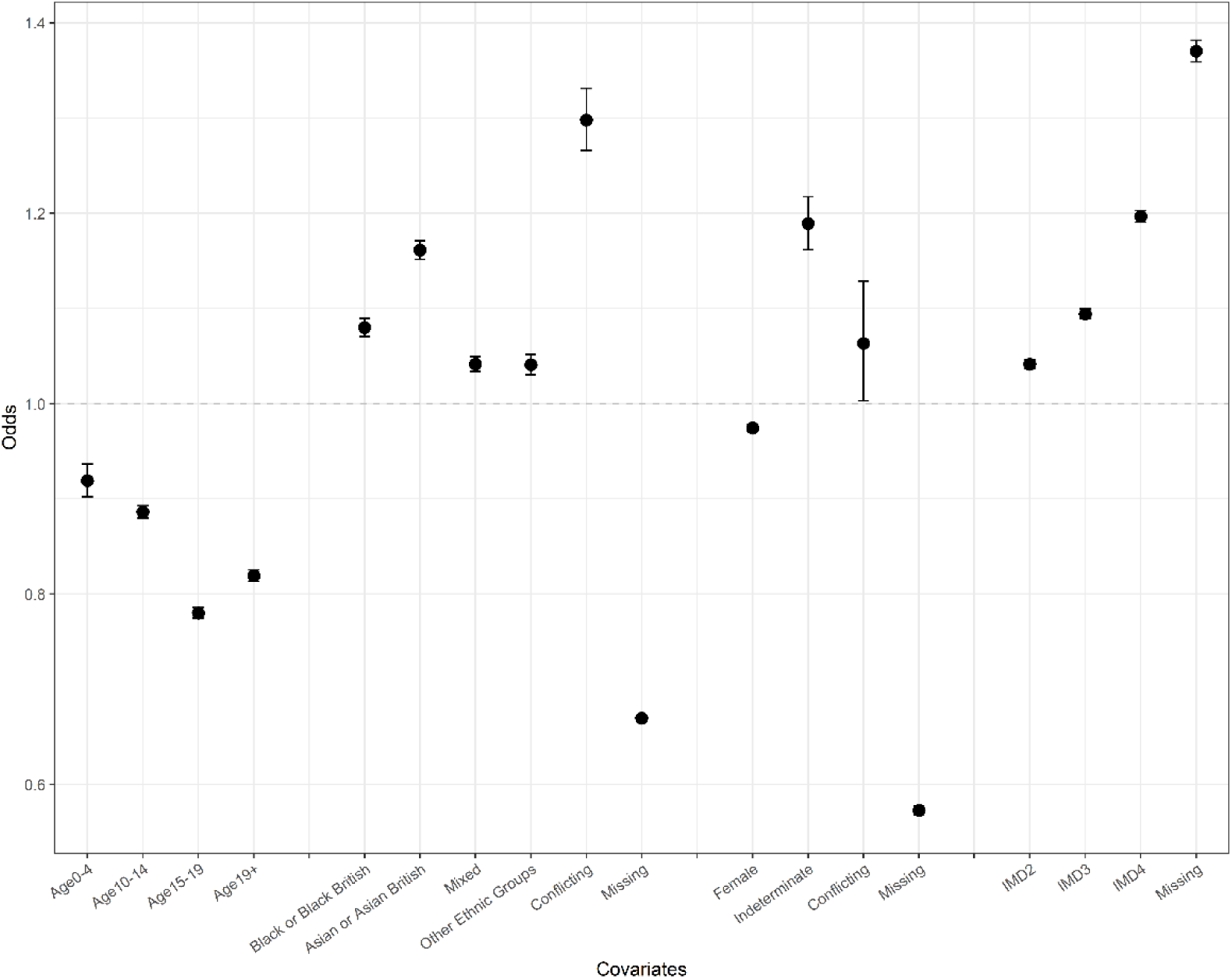
Estimated odds ratios of appointment attendance from the regression model treating missing and conflicting data as additional categories adjusted for age gender ethnicity and IMD

### Spatial and temporal patterns of service use

The percentage of CYP in contact with mental health services varied across England, ranging from 1.7% (n=8,339) in West Somerset to 7.5% (n=25,234) in South Tyneside over the eight-year period, relative to the total CYP population in each local authority (Figure 3). In 2023, local authorities where more than 10% of CYP were in contact with mental health services were predominately clustered in North East, South East, and South West regions. Absolute numbers of CYP in contact with services are shown in supplemental Figure. When considering care contact attendance, i.e., CYP who attended at least one appointment out of those referred to or in contact with services, the proportions also varied geographically, from 39% (n=18,151) in Coventry to 82% (n=16,428) in Middlesbrough (Supplemental Figure 3), with higher attendance generally in the North and Midlands.

**Figure 3.**
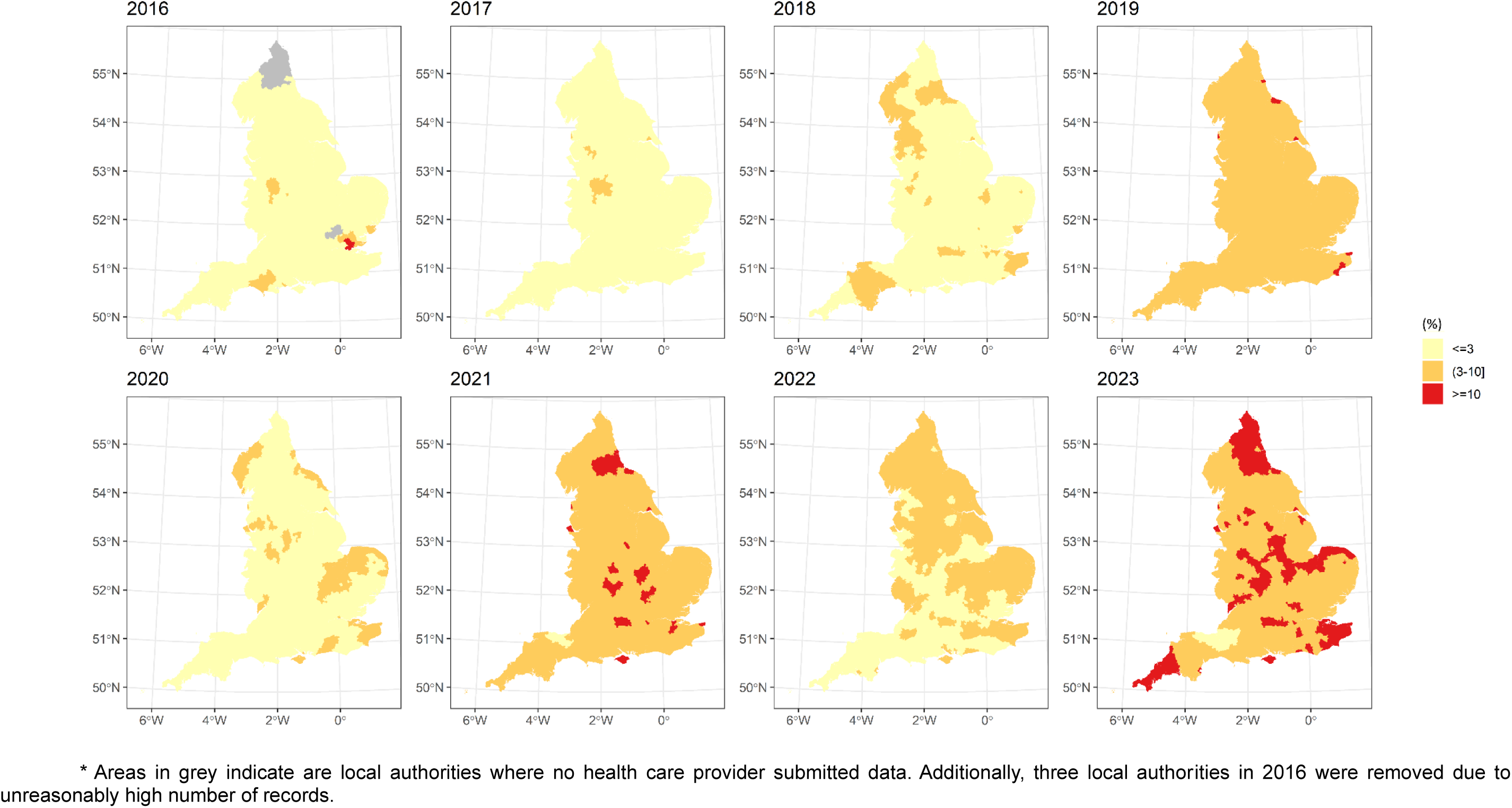
Local authority variation in the percentage of children and young people in contact with mental health services (referred to mental health services) in England from financial years 2016 to 2023.

A subset of CYP had recorded contact with services but no attended care contact (unlinked cases). The spatial distribution of these unlinked cases remained stable across years, with only a few local authorities showing consistently high proportions (supplemental Figure 4).

Nationally, the proportion of CYP in contact with services increased from 1.9% (n=371,655) in 2016 to 9.7% (n=1,699,899) in 2023 (solid line in Figure 4). The proportion attending at least one care contact peaked at 65% (n=767,079) in 2019 and then stabilised around 60% (n=1,016,976) by 2023 (dashed line in Figure 4). Monthly submission patterns revealed substantial surges near the end of each financial year, particularly after 2019, likely reflecting reporting and administrative cycles (Supplemental Figure 5).

**Figure 4.**
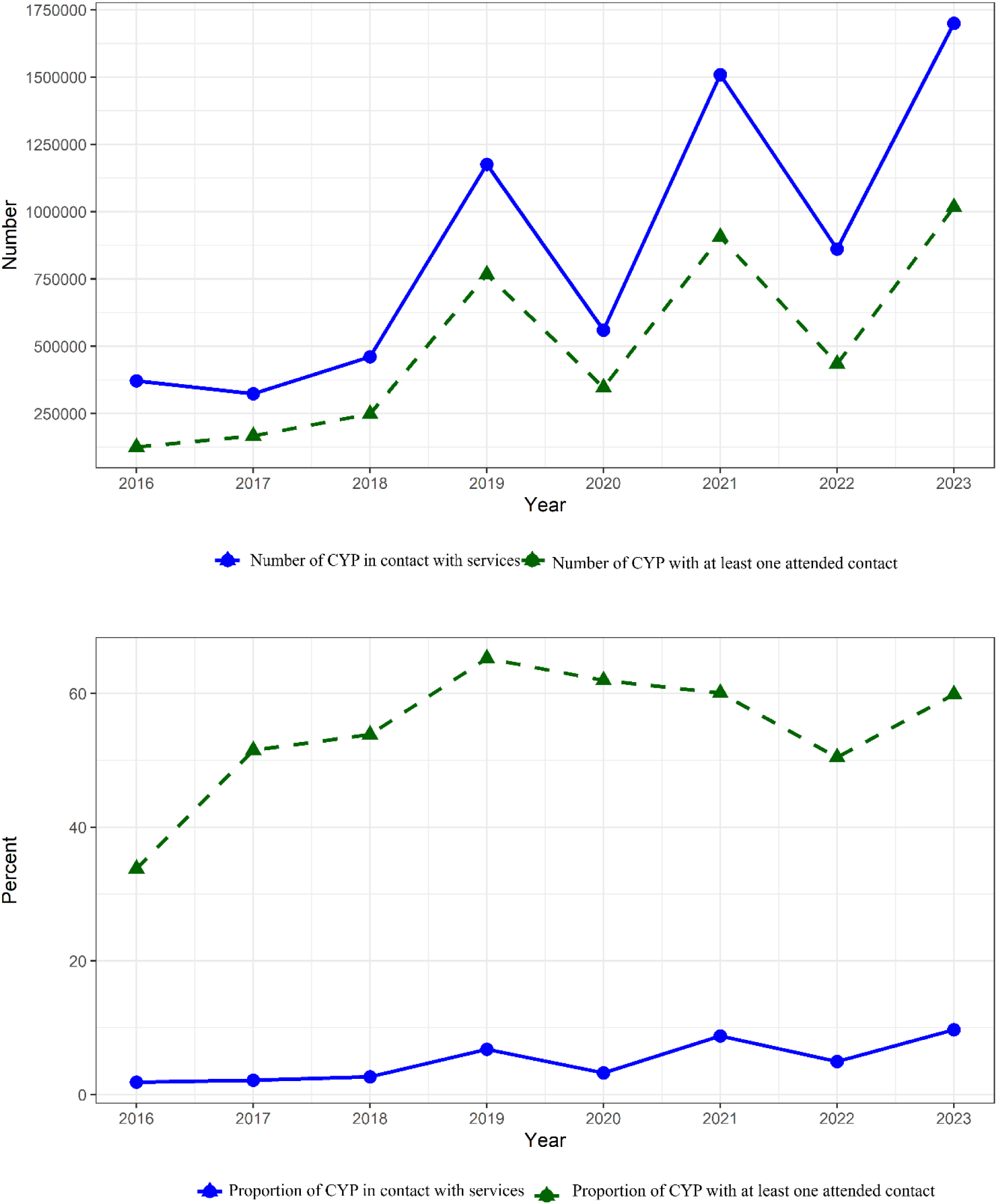
Temporal trend of the proportion and number of CYP in contact with mental health service and have at least one attended care contact

### Sociodemographic profiling

Detailed sociodemographic patterns are detailed in Supplementary Tables 1 and 2. Until 2019, care contact attendance rates were similar for males and females, but from 2020 onwards, females consistently attended at higher rates. Attendance was highest among 15– 19-year-olds, while rates among 10–14-year-olds increased and became comparable after 2020.

### Sensitivity analysis

Restricting analyses to 77 providers that submitted records consistently from 2016 to 2023 did not materially change spatial or temporal trends (supplemental Figures 5-7). Re-estimating the regression coefficients for 2021 data produced comparable results, except that the odds of attendance for CYP with missing gender data differed between 2023 (OR=0.57, 95% CI: 0.56–0.58) and 2021 (OR=1.26, 95% CI: 1.25–1.27).

## Discussion

Between 2016 and 2023, nearly 5 million CYP in England had some form of contact with mental health services, with 62% attending at least one care contact. In 2023, 9.7% of CYP were in contact with mental health services, and 60% attended at least one care contact. These patterns remained robust when analyses were limited to consistently reporting providers, suggesting that observed regional and temporal trends reflect real variation rather than artefacts of reporting.

### Data completeness and consistency

The MHSDS is a unique national resource for monitoring mental-health service use and guiding resource allocation. Despite its potential, the MHSDS remains underutilised in research, partly due to the inherent data quality challenges associated with administrative datasets. Although data quality has improved over the years, substantial missingness persists, especially for ethnicity, geographical location, and socioeconomic status. Missingness was systematic and often was linked to specific subpopulations. For example, CYP with missing ethnicity data were more likely to have missing gender information and lower appointment attendance, suggesting non-random patterns. Regression models showed that CYP with missing gender or ethnicity had lower odds of attending care contacts compared to those with known data. In contrast, missing IMD was associated with higher attendance. However, around 70% of CYP with missing IMD were linked to non-NHS or independent providers, including online services, and excluding these reverse the association. Although confidence intervals for “missing” categories were narrow, this likely reflects the large sample and internal homogeneity of these records.

While most records were internally consistent, a small proportion had conflicting gender data, raising questions about whether this discrepancy reflected data entry errors or real changes in identity. Although these cases were rare, they highlight the importance of temporal tracking and validation of sociodemographic fields within administrative datasets.

Excluding incomplete cases removed one-third of observations, which could introduce bias and reduce statistical power. Including missing categories preserved data but introduced residual confounding ^16^. Methods like Multiple Imputation (MI), assuming Missing at Random (MAR), may offer a better solution, especially if missing data is due to provider-level reporting. Sensitivity analyses showed that missingness effect varied by year, likely reflecting evolving data collection practices.

This study highlights the dataset’s strengths, identifies areas for improvement, and provides practical recommendations, for both NHS England and for researchers, to enhance its utility for future research and policymaking (Box 1).

### Box 1: Enhancing research utility of the MHSDS

1. Enhance metadata transparency and data cleaning workflows

There is a need for clearer documentation on the derivation and evolution of key variables (e.g., ethnicity, IMD, referral pathways) to enhance reproducibility. Light-touch, automated data validation at source, such as removing duplicates and flagging potential outliers, could further improve data quality before extracts are released to researchers. Providing a set of data quality notes would help researchers interpret data anomalies (e.g., unusually high numbers of CYP in contact with mental health services in some local authorities in 2016).

2. Establish standardised guidelines for cross-sectional and longitudinal data use Establishing consistent linkage keys and table structures across years would strengthen the reproducibility of longitudinal analyses. Clear methodological guidance for merging tables and addressing changes over time could reduce analytical variability between studies.

3. Address systematic missingness

Improving data collection, linkage, and imputation strategies are needed to reduce gaps, particularly for key variables like gender identity, ethnicity, and socioeconomic status (e.g., IMD deciles). More granular data on these factors would improve analyses and reduce biases.

4. Ensuring consistent geographic resolution

The spatial resolution of data has varied over time, limiting consistency, particularly in earlier years. Future data collection should aim for consistent geographic resolution (e.g., LSOA data for all years) to support more accurate spatial analyses.

5. Accounting for submission timing effects

Variations in data submission timing can affect service access patterns, particularly around financial year-end reporting deadlines. Including metadata on submission timing and providing analytical guidance to adjust for this effect would improve interpretability of analyses.

6. Recognise service-specific data characteristics

Our analysis showed that independent and non-NHS providers contributed to high proportion of missingness. Excluding these providers led to slight changes in model coefficients, reflecting their unique demographic profiles. Understanding the context of data from different service providers is essential for accurate analysis and interpretation.

7. Strengthening collaboration between data custodians and researchers

Creating mechanisms, such as structured feedback channels, workshops, or discussion forums, for researchers to report recurring data quality issues and ask questions could foster iterative improvement in data quality without adding burdens to frontline services

## Spatial, temporal, and sociodemographic patterns

Spatially, northern and midlands regions showed higher attendance rates, while southern and eastern areas had higher contact but lower attendance, suggesting regional differences in service capacity, policies, or access barriers. Further studies are needed to understand the underlying drivers of these variations. Over time, the number of CYP in contact with mental health services steadily increased, but the proportion attending at least one care contact has remained relatively static since 2019, indicating rising demand without proportional service expansion. This aligns with previous reports highlighting funding gaps and staff shortages in mental health care delivery ^17^. Temporary dips in 2020 and 2022 correspond to COVID-19 disruption and 2022–23 NHS cyber incident.

Females aged 15–19, particularly from deprived areas, had the highest attendance, suggesting elevated mental-health burden in this group. Attendance among 10–14-years-olds increased over time, potentially reflecting earlier intervention initiatives^3^. Inclusion of infants (<1 year) likely reflects parental referrals rather than direct child presentation^18^.

Before the COVID-19 pandemic, gender difference in attendance were minimal. However, post-pandemic, females accounted for most attendances, which aligns with reports of increased mental health challenges among adolescent girls during the pandemic ^19^. These challenges have been postulated to be driven by factors such as social isolation and increased social media use ^20^ ^21^.

## Strength and limitation

This study demonstrates the value and challenges of using administrative datasets for mental health research. Analyses of temporal, spatial, and sociodemographic patterns revealed persistent inequalities and data quality issues that need be addressed to ensure equitable service provision.

Our study has some limitation. We analysed each financial year of data separately, which may misclassify ongoing treatment episodes. For example, CYP who are in contact with mental health services late in one year and continue treatment into the following year may be classified as not receiving treatment in the first year, even though they were in ongoing care. Longitudinal linkage within MHSDS not only allows CYP to be followed over time to derive treatment journeys but can also serve to reduce missingness using longitudinal imputation of variables such as ethnicity gender LSOA and IMD. This study aimed to establish baseline patterns of being in contact with mental health services and care contact attendance rather than describing longitudinal care trajectories. While this research examined who is in contact with mental health services and begins engagement by attending appointments, a broader investigation is needed to understand diagnostic and treatment pathways to assess the accuracy and consistency of diagnosis records.

## Clinical implications

The MHSDS is a powerful tool for evaluating national mental-health services for CYP. Improving completeness, consistency, and transparency of core sociodemographic variables is essential to generate reliable and equitable evidence. Data incompleteness can influence analyses, emphasizing the need for improved data collection practices and the adoption of robust methods to address missing values. This ensures delivering equitable, data-driven mental-health care for children and young people across England.

## Data Availability

Data can be obtained by submitting a request to the Data Access Request Service (DARS). For any data enquiries contact the Enquiries team by emailing

## Acknowledgements

This research benefits from and contributes to the NIHR Children and Families Policy Research Unit (NIHR206114) but was not commissioned by the National Institute for Health and Care Research (NIHR) Policy Research Programme. The views expressed are those of the author(s) and not necessarily those of the NIHR or the Department of Health and Social Care.

