## Supplemental material for "Using the English national health service dataset for research into mental health service use among children and young people"

### Details of data structure and Integration in MHSDS

Each table of MHSDS is structured with unique identifiers (primary keys) and reference points (foreign keys) for data integration^1^. For instance, the Person_ID field serves as a primary key in the patient demographics table, uniquely identifying individuals, and also as a foreign key in related tables such as care contacts and referrals. This relational structure enables data linkage across tables, providing a comprehensive view of an individual’s interactions with mental health services.

For this study, we used the Master Patient Index (MHS001) and Care Contacts (MHS201) tables for the financial years 2016-2023, inclusive. The MHS001 table contains information on sociodemographic information, geographical information of residence. It includes all individuals referred to NHS-commissioned mental health, learning disabilities, or autism services, irrespective of whether they go on to have a care contact.

The MHS201 table includes data on care contacts, which refer to any interaction between patients and mental health services such as appointments, outpatient visits, telephone consultations, or follow-up meetings. It provides details on the date, commissioning health care provider, attendance, communication method, and the location of contact.

The number of CYP in contact with services was calculated as the count of those with at least one record in MHS001. The appointment attendance was determined by linking the MHS001 and MHS201 tables using a unique ID for each individual, created from a combination of individual and health provider codes, specifically “Person_ID” and “Origidprov”. To avoid duplication, we included only one record per individual per health care provider, selecting the most recent entry. The number of CYP who attended at least one care contact was calculated as the count of those with linked records in MHS001 and MHS201 and flagged as having attended the care contact.

### Details of covariates in MHSDS

**Age Calculation**

Age was calculated by subtracting the individual’s date of birth from the first day of the month in which the data were submitted. For analysis, ages were grouped into the following categories: 0–4, 5–9, 10–14, 15–19, and 20–25 years.

**Ethnicity Data Processing**

In the MHSDS, ethnicity is recorded using three variables: NHS ethnicity code, high-level ethnicity group, and low-level ethnicity group.

•	NHS ethnicity codes: These group ethnicity into 16+1 categories, which have been revised with each census, resulting in differing code notations based on the census data entry.

•	High-level ethnicity groups: Based on ONS ethnicity classification, these include Asian/Asian British, Black/Africa/Caribbean,/Black British, Mixed, Other Ethnic groups, Unknown, and White.

•	Low level ethnicity groups: Based on ONS ethnicity classification, these include British, Irish, Traveller, any other White background, White and Black Caribbean, White and Black African, White and Asian, any other Mixed background, Indian, Pakistani, Bangladeshi, Chinese, any other Asian background, African, Caribbean, any other Black background, Arab, Any other ethnic group, not stated, and unknown.

For consistency, high-level classifications were used, including White, Black or Black British, Asian or Asian British, Mixed, and Other Ethnic Groups. A validated algorithm by Pineda-Moncusi et al. ^2^was applied to map NHS ethnicity codes to these categories when inconsistencies arose due to changes in recording practices.

**Handling Missing or Inconsistent Gender and Ethnicity Records**

Gender and ethnicity are ideally self-declared^3^ but may vary across records due to updates during care episodes. We used a gender or ethnicity code if it appeared at least once across an individual's records and had a meaningful value. If no valid code was present, we labelled them as missing. Some individuals had codes indicating “not stated” or “unknown recorded”. “Not stated” applied to individuals who declined to provide information, while “unknown” referred to not known information. If there were multiple inconsistent characteristics for an individual CYP (e.g., gender labelled as female in one record and male in another), these were labelled as “conflicting” records. While “conflicting” term might also capture identity complexities, defining and analysing these conflicts helps assess their frequency and impact. Misspecified codes (e.g., N, F1, etc rather than M or F for male or female) were treated as missing.

**Index of Multiple Deprivation (IMD)**

The IMD measures relative deprivation across small areas (LSOAs) using seven domains: income, employment, health and disability, education, skills and training, barriers to housing and services, living environment, and crime^4^. In the MHSDS, IMD is reported in quartiles, with the lowest quartile indicating the most deprived areas. IMD data has been available from 2019 onwards.

### Details on data preparation for the regression model

To conduct the regression model, we prepared the data so that each individual was associated with a single value for each covariate. However, since individuals may have multiple records with varying covariate values, we applied the following approach:

- We used a gender or ethnicity code if it appeared at least once across an individual's records and had a meaningful value. If no valid code was present, the value was labelled as missing.
- If an individual had inconsistent covariate values (e.g., gender recorded as female in one record and male in another), the value was labelled as conflicting.
- For neighbourhood-level socioeconomic status (measured by the Index of Multiple Deprivation [IMD]), multiple records could exist due to changes in residential address. In cases of conflicting IMD values, we assigned the value associated with the most recent care contact date.

### Table 1. Sociodemographic characteristics of children and young people in contact with mental health services between financials years 2016 and 2023

| N (%) | 2016 | 2017 | 2018 | 2019 | 2020 | 2021 | 2022 | 2023 |
| --- | --- | --- | --- | --- | --- | --- | --- | --- |
| Number in contact with mental health services | 371,655 | 323,641 | 461,081 | 1,175,705 | 559,616 | 1,508,842 | 860,905 | 1,699,899 |
| Gender | | | | | | | | |
| Male | 183,507 (49.38) | 163,158 (50.41) | 218,683 (47.43) | 557,362 (47.41) | 232,855 (41.61) | 643,238 (42.63) | 316,879 (36.81) | 750,013 (44.1) |
| Female | 186,438 (50.16) | 158,460 (48.96) | 227,635 (49.37) | 570,156 (48.49) | 299,608 (53.54) | 782,924 (51.89) | 475,055 (55.2) | 862,739 (50.7) |
| Missing | 870  (<1) | 1,675  (<1) | 13,396  (2.9) | 42,573  (3.6) | 21,682  (3.9) | 64,505  (4.3) | 49,261  (5.7) | 74,539 (4.4) |
| Indeterminate | 775  (<1) | 228  (<1) | 1,232  (<1) | 4,835  (<1) | 5,126  (<1) | 17,463  (1.2) | 19,507  (2.3) | 11,780  (<1) |
| Conflicting | 65 (0<1) | 120 (<1) | 134 (<1) | 774 (<1) | 345 (<1) | 712 (<1) | 203 (<1) | 818 (<1) |
| Ethnicity | | | | | | | | |
| White | 371,655 (55.5) | 172,046 (53.2) | 252,716 (54.8) | 709,624 (60.4) | 322,217 (57.6) | 709,042 (47.0) | 519,708 (60.4) | 1,000,733 (58.9) |
| Asian | 206,429 (3.2) | 11,542  (3.6) | 16,459  (3.6) | 44,170  (3.8) | 21,783  (3.9) | 50,581  (3.3) | 43,711  (5.0) | 75,052  (4.4) |
| Black | 1,180 (3.8) | 8,908 (2.7) | 11,485 (2.5) | 31,788 (2.7) | 14,514 (2.6) | 34,269 (2.3) | 25,943 (3.0) | 51,189  (3.0) |
| Mixed | 9,502 (2.6) | 9,552 (3.0) | 14,805 (3.2) | 46,012 (3.9) | 20,904 (3.7) | 53,176 (3.5) | 41,690 (4.8) | 84,105 (4.9) |
| Other | 8,633 (2.3) | 9,129 (2.8) | 11,481 (2.5) | 23,448 (2.0) | 11,611 (2.0) | 27,517 (1.8) | 21,705 (2.5) | 45,526(2.7) |
| Conflicting | 608 (<1) | 391 (<1) | 640 (<1) | 2,866 (<1) | 837 (<1) | 13,872 (<1) | 1,431 (<1) | 37,210 (2.2) |
| Missing | 120,452 (32.4) | 112,073 (34.6) | 153,495 (33.3) | 317,797 (27.0) | 167,750 (30.0) | 620,385 (41.1) | 206,717 (24.0) | 406,084 (23.9) |
| IMD | | | | | | | | |
| 1  (most deprived) | - | - | - | 363,521 (30.6) | 147,896 (26.1) | 438,878 (28.9) | 197,816 (22.9) | 519,808 (30.3) |
| 2 | - | - | - | 268,263 (22.6) | 110,880 (19.7) | 328,324 (21.6) | 146,095 (16.9) | 389,733 (22.8) |
| 3 | - | - | - | 216,579 (18.2) | 91,081 (16.2) | 269,459 (17.7) | 122,079 (14.1) | 320,438 (18.7) |
| 4 (least deprived) | - | - | - | 192,145 (16.2) | 81,011 (14.4) | 243,067 (16.0) | 107,672 (12.5) | 287,691 (16.8) |
| Missing | 100% | 100% | 100% | 146,035 (12.3) | 131,312 (23.4) | 240,573 (15.8) | 290,746 (33.6) | 194,868 (11.4) |
| Age | | | | | | | | |
| 0-4 | 31,251 (8.6) | 9,640 (3.0) | 10,461 (2.3) | 26,618 (2.3) | 13,678 (2.4) | 31,051 (2.0) | 13,242 (1.5) | 41,279 (2.4) |
| 5-9 | 56,130 (15.4) | 43,126 (13.4) | 59,561 (12.9) | 166,755 (14.2) | 64,992 (11.6) | 197,551 (13.) | 90,805 (10.5) | 265,535 (15.6) |
| 10-14 | 71,408 (19.6) | 69,286 (21.5) | 116,478 (25.3) | 345,056 (29.3) | 153,781 (27.5) | 478,730 (31.6) | 293,817 (34.1) | 529,227 (31.1) |
| 15-19 | 116,996 (32.1) | 111,629 (34.6) | 156,583 (34.0) | 378,676 (32.2) | 206,650 (37.0) | 502,675 (33.2) | 314,473 (36.5) | 524,890 (30.8) |
| >19 | 88,858 (24.4) | 89,010 (27.6) | 117,208 (25.5) | 260,181 (22.1) | 119,426 (21.4) | 302,723 (20.0) | 148,375 (17.2) | 341,123 (20.0) |
| IMD: Index of Multiple Deprivation | | | | | | | | |

### Table 2 Sociodemographic characteristics of children and young people with at least one attended care contact between financial years 2016 and 2023

|  | 2016 | 2017 | 2018 | 2019 | 2020 | 2021 | 2022 | 2023 |
| --- | --- | --- | --- | --- | --- | --- | --- | --- |
| Number with at least one attended care contact | 125,611 | 166,711 | 248,320 | 767,079 | 346,879 | 906,426 | 434,306 | 1,016,976 |
| Gender | | | | | | | | |
| Male | 60,164 (47.9%) | 81,394 (48.8%) | 112,141 (45.2%) | 346,002 (45.1%) | 130,293  (37.6%) | 374,496 (41.3%) | 153,121 (35.3%) | 415,024 (40.8%) |
| Female | 64,502 (51.3%) | 84,116 (50.5%) | 131,138 (52.8%) | 394,112 (51.4%) | 203,088 (58.5%) | 483,266 (53.3%) | 242,888 (55.9%) | 543,207 (53.4%) |
| Indeterminate | 559  (<1%) | 113  (<1%) | 1,065 (<1%) | 4,322 (<1%) | 4,739 (1.4%) | 8,674  (<1%) | 8,525 (2.0%) | 9,693  (<1%) |
| Conflicting | 21  (<1%) | 40  (<1%) | 44  (<1%) | 321 (<1%) | 103  (<1%) | 287  (<1%) | 84  (<1%) | 322  (<1%) |
| Missing | 365  (<1%) | 1,048 (<1%) | 3,932 (1.6%) | 22,322 (2.9%) | 8,656 (2.5%) | 39,703 (4.4%) | 29,649 (6.8%) | 48,711 (4.8%) |
| Ethnicity | | | | | | | | |
| White | 76,938 (61.2%) | 97,669 (58.6%) | 150,285 (60.5%) | 498,421  (65.0%) | 218,521 (63.0%) | 455,661  (50.3%) | 270,589 (62.3%) | 658,369 (64.7%) |
| Asian | 4,312 (3.4%) | 5,845 (3.5%) | 9,360 (3.8%) | 31,264  (4.1%) | 15,453 (4.4%) | 30,478  (3.4%) | 22,264 (5.1%) | 48,712 (4.8%) |
| Black | 3,446 (2.7%) | 4,532 (2.7%) | 6,624 (2.7%) | 22,565 (2.9%) | 9,988 (2.9%) | 21,628 (2.4%) | 13,131 (3.0%) | 33,878 (3.3%) |
| Mixed | 3,512 (2.8%) | 4,985 (3.0%) | 8,221 (3.3%) | 30,468 (4.0%) | 13,552 (3.9%) | 33,244  (3.7%) | 21,418 (4.9%) | 54,336 (5.3%) |
| Other | 3,633 (3.0%) | 4,655 (2.8%) | 6,811 (2.7%) | 15,903 (2.1%) | 7,338 (2.1%) | 16,767  (1.8%) | 10,517 (2.4%) | 29,034 (2.8%) |
| Conflicting | 423  (<1%) | 481  (<1%) | 739 (<1%) | 3,754 (<1%) | 300  (<1%) | 807  (<1%) | 460  (<1%) | 2,221  (<1%) |
| Missing | 33,347 (26.5%) | 48,544 (29.1%) | 66,280 (26.7%) | 164,704 (21.5%) | 81,727 (23.6%) | 347,841 (38.4%) | 95,927 (22.1%) | 190,426 (18.7%) |
| IMD | | | | | | | | |
| 1 (most deprived) | NA | NA | NA | 235,970 (31.0%) | 86,463 (24.9%) | 279,533 (30.7%) | 110,405 (25.3%) | 309,022 (30.2%) |
| 2 | NA | NA | NA | 172,732 (22.4%) | 64,244 (18.5%) | 207,846 (22.8%) | 82,107 (18.8%) | 228,113 (22.3%) |
| 3 | NA | NA | NA | 138,464 (18.0%) | 53,120 (15.3%) | 170,385 (18.7%) | 69,318 (15.9%) | 185,658 (18.2%) |
| 4 (least deprived) | NA | NA | NA | 120,106 (15.6%) | 46,915 (13.5%) | 151,078 (16.6%) | 59,950 (13.8%) | 163,090 (16.0%) |
| Missing | 100% | 100% | 100% | 104,182 (13.5%) | 97,112 (27.9%) | 102,355 (11.2%) | 113,774 (26.1%) | 135697 (13.3%) |
| Age | | | | | | | | |
| 0-4 | 2,796 (2.2%) | 4,057 (2.4%) | 4,277 (1.7%) | 11,539 (1.5%) | 4,814 (1.4%) | 12,896 (1.42%) | 5,189 (1.2%) | 15,111 (1.5%) |
| 5-9 | 11,790 (9.4%) | 18,262 (11.0%) | 25,541 (10.3%) | 84,435 (11.0%) | 28,763 (8.3%) | 97,785 (10.8%) | 40,146 (9.2%) | 122,290 (12.0%) |
| 10-14 | 21,326 (17.1%) | 34,360 (20.7%) | 62,787 (25.3%) | 230,405 (30.0%) | 99,898 (28.9%) | 282,841 (31.2%) | 145,270 (33.5%) | 332,045 (32.7%) |
| 15-19 | 47,017 (37.6%) | 60,543 (36.4%) | 90,289 (36.4%) | 266,160 (34.8%) | 139,206 (40.2%) | 321,193 (35.4%) | 162,698 (37.5%) | 348,586 (34.3%) |
| 19+ | 42,029 (33.6%) | 48,896 (29.4%) | 64,796 (26.2%) | 174,527 (22.7%) | 73,406 (21.2%) | 192,738 (21.2%) | 80,610 (18.6%) | 198,702 (19.5%) |
| IMD: Index of Multiple Deprivation | | | | | | | | |

### Table 3. Sociodemographic characteristics of children and young people with at least one attended care contact with missing information on gender, ethnicity, and lower super output area of residence between financials years 2016 and 2023

| N () | 2016 | 2017 | 2018 | 2019 | 2020 | 2021 | 2022 | 2023 |
| --- | --- | --- | --- | --- | --- | --- | --- | --- |
| Number (percent) with missing gender | 194  (<1) | 820  (<1) | 3351 (1.35) | 6970  (<1) | 3,864  (1.1) | 27,306 (3.01) | 16,723 (3.85) | 23,142 (2.3) |
| Number (percent) with missing ethnicity | 33,347 (26.5) | 48,544 (29.1) | 66,280 (26.7) | 164,704 (21.5) | 83,679  (24.0) | 347,841 (38.4) | 95,927 (22.1) | 190,426 (18.7) |
| Number (percent) with missing LSOA | 125,611 (100) | 5,454  (3.3) | 28,986 (11.7) | 100,284 (13.1) | 95,903 (27.6) | 98,004 (10.8) | 111,943 (25.8) | 131,911 (13.0) |
| Characteristics of CYP who entered treatment and lack gender information | | | | | | | | |
| *Ethnicity* | | | | | | | | |
| White | 137  (70.6) | 28  (3.4) | 15  (<1) | 12  (<1) | 141  (3.6) | 13,422 (49.1) | 6,326 (37.8) | 9,139 (39.5) |
| Asian | <10  (<1) | <10  (<1) | <10  (<1) | <10  (<1) | <10  (<1) | 928  (3.4) | 926  (5.5) | 1,007  (4.3) |
| Black | <10 (<1) | <10 (<1) | <10 (<1) | <10 (<1) | <10 (<1) | 339 (1.2) | 128 (<1) | 439 (1.9) |
| Mixed | <10 (<1) | <10 (<1) | <10 (<1) | <10 (<1) | 15 (<1) | 1,302 (4.8) | 370 (2.2) | 781 (3.4) |
| Other | <10(<1) | <10(<1) | <10 (<1) | <10 (<1) | <10 (<1) | 281 (1.0) | 189 (1.1) | 256 (1.1) |
| Conflicting | - | - | - | - | - | 1 (<1) | 5 (<1) | - |
| Missing | 54  (27.8) | 790  (96.3) | 3,330 (99.4) | 6,952  (99.7) | 3,772  (95.6) | 11,033 (40.4) | 8,779 (52.5) | 11,520 (49.8) |
| *IMD* | | | | | | | | |
| 1 | - | - | - | 2,055  (29.5) | 931  (23.6) | 98,929 (28.4) | 5,091 (30.4) | 7,225 (31.2) |
| 2 | - | - | - | 923  (13.2) | 430  (10.9) | 76,652 (22.0) | 2,938 (17.6) | 4,745 (20.5) |
| 3 | - | - | - | 387  (5.5) | 236  (6.0) | 65,297 (18.8) | 2,586 (15.5) | 4,526 (19.6) |
| 4 | - | - | - | 346  (5.0) | 194  (4.9) | 59,142 (17.0) | 2,525 (15.1) | 4,233 (18.3) |
| Missing | 194  (100) | 820  (100) | 3,351 (100) | 3,259  (46.7) | 2,156  (54.6) | 48,892 (14.1) | 3,583 (21.4) | 2,413 (10.4) |
| *Age* | | | | | | | | |
| 0-4 | 12  (6) | 0 (0) | <10 | 14 (<1) | 48 (1.2) | 377 (1.4) | 129 (<1) | 149 (<1) |
| 5-9 |  | 62  (7.6) | 327  (9.7) | 523  (7.5) | 380  (9.9) | 3,843 (14.1) | 2,018 (12.1) | 2,827 (12.2) |
| 10-14 | 36  (18.56) | 328  (40) | 2,013 (60.1) | 4,052  (58.2) | 1,889  (49.3) | 11,868 (43.5) | 8,028 (48.1) | 9,666 (41.8) |
| 15-19 | 76  (39.2) | 303  (37.0) | 1,000 (30.0) | 2,318  (33.3) | 1,470  (38.3) | 8,674 (31.8) | 5,625 (33.7) | 7,510 (32.5) |
| >19 | 70 (36.1) | 127 (15.5) | <10 (<1) | 53 (<1) | 46 (1.2) | 2529 (9.3) | 894 (5.3) | 2967 (12.8) |
| Characteristics of CYP who entered treatment and lack ethnicity information | | | | | | | | |
| *Gender* | | | | | | | | |
| Male | 15,904 (47.7) | 23,449 (48.3) | 29,451 (44.4) | 73,576 (44.7) | 33,064 (40.4) | 137,003 (39.4) | 34,727 (36.2) | 78,721 (41.3) |
| Female | 17,191 (51.5) | 24,150 (49.7) | 33,104 (49.9) | 78,372 (47.6) | 41,015 (50.2) | 189,699 (54.5) | 46,462 (48.4) | 92,100 (48.4) |
| Missing | 135  (<1) | 907  (1.9) | 3,574  (5.3) | 12,172  (7.4) | 7,184  (8.8) | 17,310 (5.0) | 8,779 (9.1) | 18,088 (9.5) |
| Indeterminate | 110  (<1) | 28  (<1) | 140  (<1) | 567  (<1) | 453  (<1) | 3,765  (1.1) | 1,237 (1.29) | 1,482  (<1) |
| Conflicting | <10  (<1) | 10  (<1) | <10  (<1) | 17  (<1) | 11  (<1) | 64  (<1) | 17  (<1) | 34  (<1) |
| *IMD* | | | | | | | | |
| 1 | NA | NA | NA | 48,252 (29.3) | 22,261 (27.2) | 98,929 (28.4) | 26,305 (27.4) | 56,363 (29.6) |
| 2 | NA | NA | NA | 38,528 (23.4) | 18,302 (22.4) | 76,652 (22.0) | 20,504 (21.4) | 44,058 (23.1) |
| 3 | NA | NA | NA | 33,075 (20.1) | 15,942 (19.5) | 65,297 (18.8) | 18,162 (18.9) | 38,192 (20.1) |
| 4 | NA | NA | NA | 29,522  (17.9) | 14,954 (18.3) | 59,142 (17.0) | 16,928 (17.6) | 35,037 (18.4) |
| Missing | 33,347 (100) | 48,544 (100) | 66,280 (100) | 15,655  (9.5) | 10,381 (12.7) | 48,892 (14.0) | 14,163 (14.8) | 17,139 (9.0) |
| *Age* | | | | | | | | |
| 0-4 | 867  (2.6) | 1,551  (3.2) | 1,224  (1.8) | 2,783  (1.7) | 1,391  (1.7) | 5,229  (1.5) | 1,713 (1.8) | 3,612  (1.9) |
| 5-9 | 3,563 (10.8) | 5,969 (12.3) | 7,723 (11.7) | 20,537 (12.5) | 8,874  (10.9) | 36,747 (10.6) | 10,615 (11.1) | 23,243 (12.3) |
| 10-14 | 6,129 (18.5) | 10,637 (22.0) | 17,130 (25.9) | 49,573 (30.1) | 21,698 (26.7) | 110,281 (31.7) | 30,254 (31.6) | 61,322 (32.4) |
| 15-19 | 12,152 (36.7) | 16,959 (35.1) | 23,647 (35.8) | 55,691 (33.9) | 29,705 (36.5) | 123,431 (35.5) | 33,205 (34.7) | 61,823 (32.7) |
| >19 | 10,408 (31.4) | 13,217 (27.3) | 16,373 (24.8) | 35,873 (21.8) | 19,647 (24.2) | 71,979 (20.7) | 19,800 (20.7) | 39,221 (20.7) |
| Characteristics of CYP who entered treatment and lack LSOA information | | | | | | | | |
| *Gender* | | | | | | | | |
| Male | 60,164 (47.9) | 2,451 (44.9) | 7,250 (25.0) | 21,461 (21.4) | 16,645 (17.3) | 19,692 (20.1) | 23,587 (21.1) | 26,022 (19.7) |
| Female | 64,502 (51.3) | 2,286 (41.9) | 19,220 (66.3) | 72,120 (71.9) | 72,371 (75.5) | 67,229 (68.6) | 75,823 (67.7) | 91,367 (69.3) |
| Missing | 365  (<1) | 712  (13.0) | 1,696  (5.8) | 3,392  (3.4) | 2,599  (2.6) | 4,208  (4.3) | 4,867 (4.3) | 7,376  (5.6) |
| Indeterminate | 559  (<1) | <10  (<1) | 820  (2.8) | 3311  (3.3) | 4,288  (4.5) | 6,875  (7.0) | 7,653 (6.8) | 7,150  (5.4) |
| Conflicting | 21  (<1) | <10  (<1) | - | - | - | - | 13  (<1) | <10  (<1) |
| *Ethnicity* | | | | | | | | |
| White | 76,938 (61.2) | 2,889 (53.0) | 18,668 (64.4) | 70,066 (69.9) | 70,693 (73.7) | 41,876 (42.7) | 79,731 (71.2) | 91,519 (69.4) |
| Asian | 4,312  (3.4) | 165  (3.0) | 1,506  (5.2) | 5,812  (5.8) | 6,107  (6.4) | 3,586  (3.7) | 7,273 (6.5) | 9,221  (7.0) |
| Black | 3,446  (2.7) | 288  (5.3) | 944  (3.3) | 3,791  (3.8) | 3,295  (3.4) | 1,914  (1.9) | 3,737 (3.3) | 5,211  (3.9) |
| Mixed | 3,512  (2.8) | 187  (4.4) | 897  (3.1) | 4,453  (4.4) | 4,392  (4.6) | 2,716  (2.8) | 5,533 (4.9) | 6,619  (5.0) |
| Other | 3,633  (2.9) | 187  (3.4) | 1,343  (4.6) | 1,287  (1.3) | 1,354  (1.4) | 1,144  (1.2) | 2,050 (1.8) | 3,358  (2.5) |
| Conflicting | 423  (<1) | <10  (<1) | <10  (<1) | <10  (<1) | <10  (<1) | <10  (<1) | <10  (<1) | <10  (<1) |
| Missing | 33,347 (26.5) | 1,740  (31.8) | 5,630 (19.4) | 14,870  (14.8) | 10,060 (10.5) | 46,770 (47.7) | 13,620 (12.2) | 15,980 (12.1) |
| *IMD* | | | | | | | | |
| 1 | -(100) | -(100) | -(100) | -(100) | -(100) | -(100) | -(100) | -(100) |
| 2 | -(100) | -(100) | -(100) | -(100) | -(100) | -(100) | -(100) | -(100) |
| 3 | -(100) | -(100) | -(100) | -(100) | -(100) | -(100) | -(100) | -(100) |
| 4 | -(100) | -(100) | -(100) | -(100) | -(100) | -(100) | -(100) | -(100) |
| Missing | -(100) | -(100) | -(100) | -(100) | -(100) | -(100) | -(100) | -(100) |
| *Age* | | | | | | | | |
| 0-4 | 2,796 (2.2) | 72 (1.3) | 100 (<1) | 342 (<1) | 194 (<1) | 212 (<1) | 86 (<1) | 110 (<1) |
| 5-9 | 11,790 (9.4) | 514  (9.5) | 796  (2.7) | 1,613  (1.6) | 893  (<1) | 2,271  (2.3) | 1,183 (1.1) | 1,633  (1.2) |
| 10-14 | 21,326  (17.1) | 1,123 (20.7) | 12,864 (44.5) | 49,119 (49.0) | 41,535 (43.3) | 44,037 (44.9) | 51,334 (45.9) | 55,396 (42.1) |
| 15-19 | 47,017 (37.6) | 1,937 (35.8) | 12,040 (41.6) | 42,452 (42.4) | 47,422 (49.5) | 43,089 (44.0) | 49,219 (44.0) | 59,304 (45.1) |
| >19 | 42,029 (33.6) | 1,766 (32.6) | 3,115 (10.8) | 6,675  (6.7) | 5,799  (6.0) | 8,353  (8.5) | 10,051 (9.0) | 15,125 (11.5) |
| IMD: Index of Multiple Deprivation  LSOA: Lower Super Output Area | | | | | | | | |

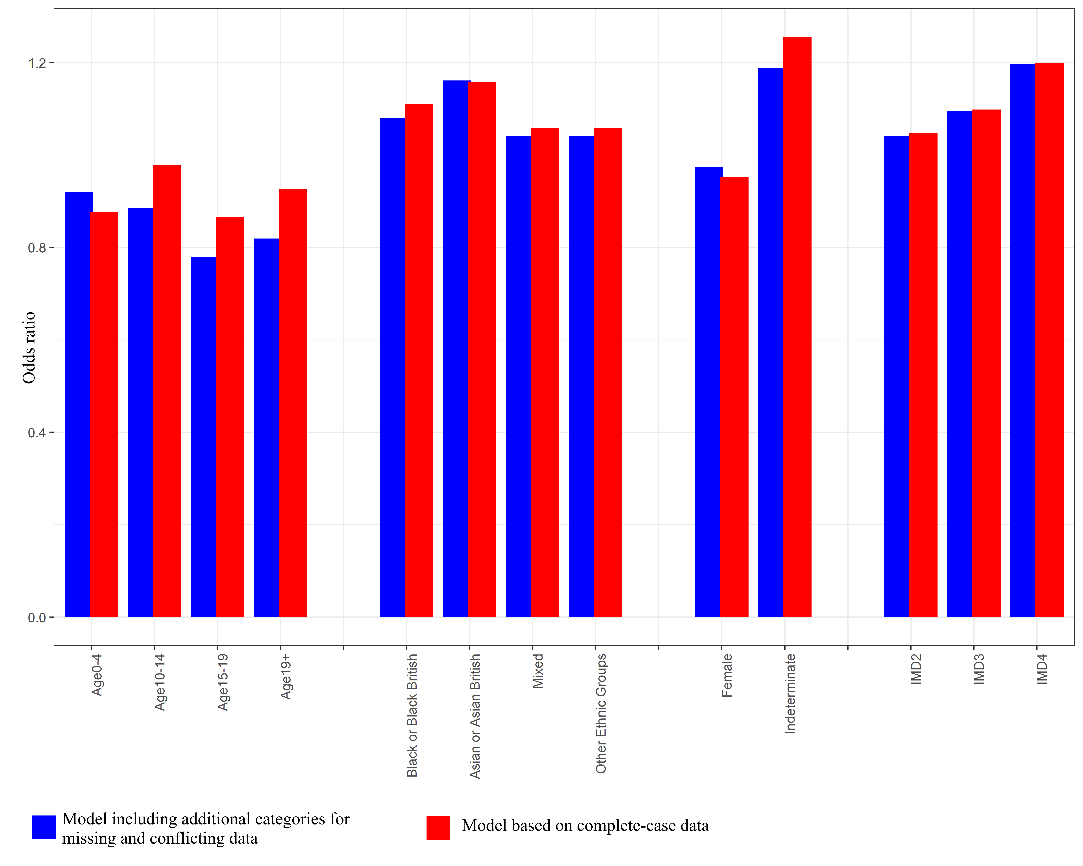

### Figure 1. Comparison of estimated odds ratios from the binomial regression model: treating missing and conflicting data as separate categories versus excluding them from the analysis for 2023 data

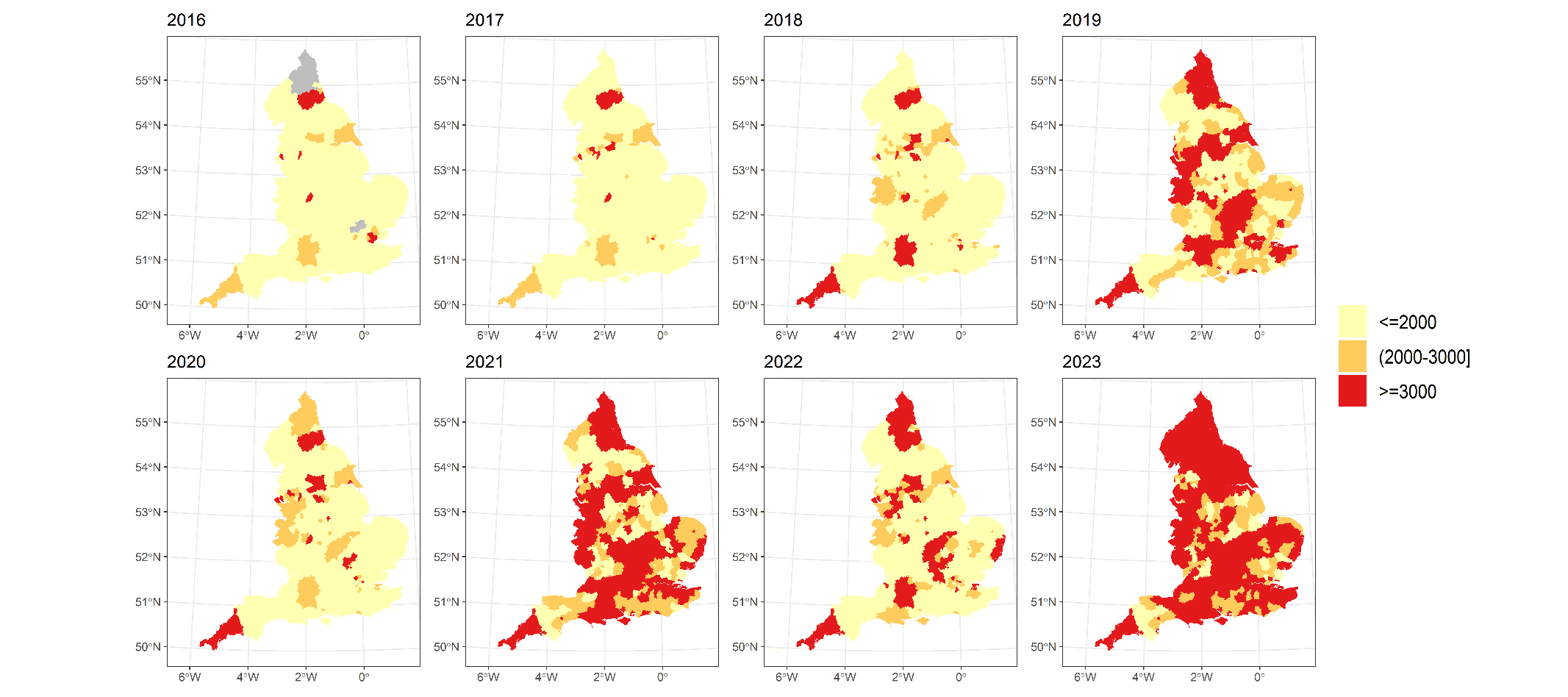

* Areas in grey indicate are local authorities where no health care provider submitted data. Additionally, three local authorities in 2016 were removed due to unreasonably high number of records.

### Figure 2. Local authority variation in the number of children and young people in contact with mental health services in England from financial years 2016 to 2023.

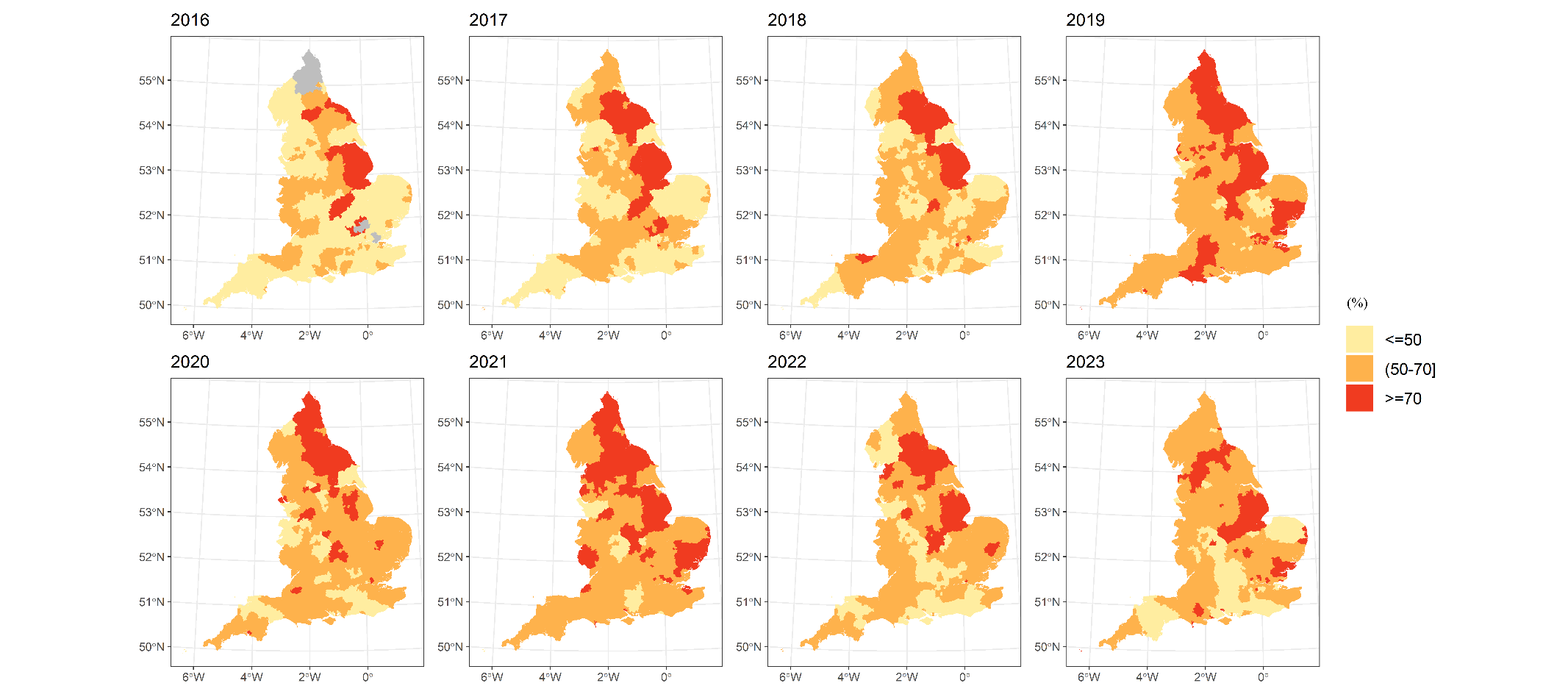
* Areas in grey indicate are local authorities where no health care provider submitted data. Additionally, three local authorities in 2016 were removed due to unreasonably high number of records.

### Figure 3. Spatial distribution of percentage of children and young people with at least one attended care contact in England from financial years 2016 to 2023.

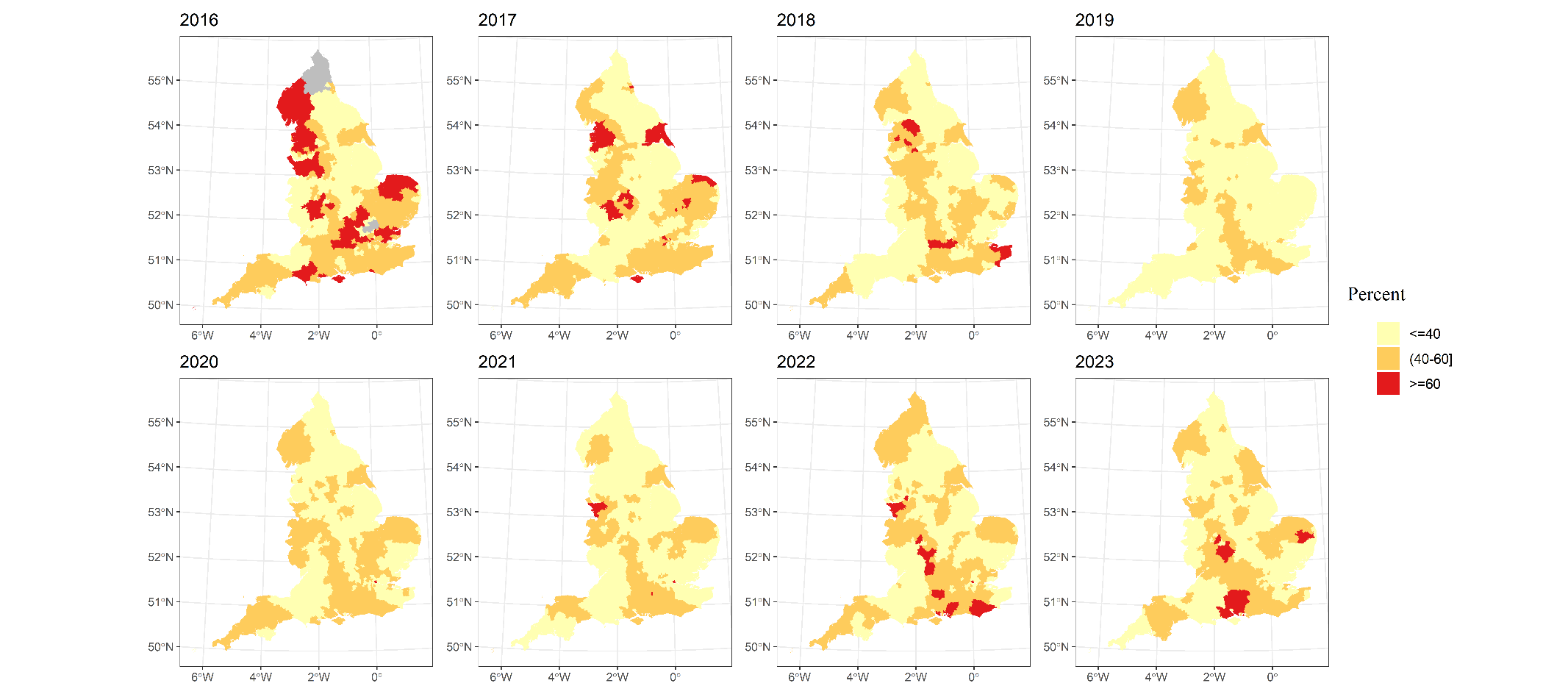

* Areas in grey indicate are local authorities where no health care provider submitted data. Additionally, three local authorities in 2016 were removed due to unreasonably high number of records.

### Figure 4. Local authority variation in the percent of children and young people who did not have any attended care contact in England from financial years 2016 to 2023. Areas in grey indicate missing data.

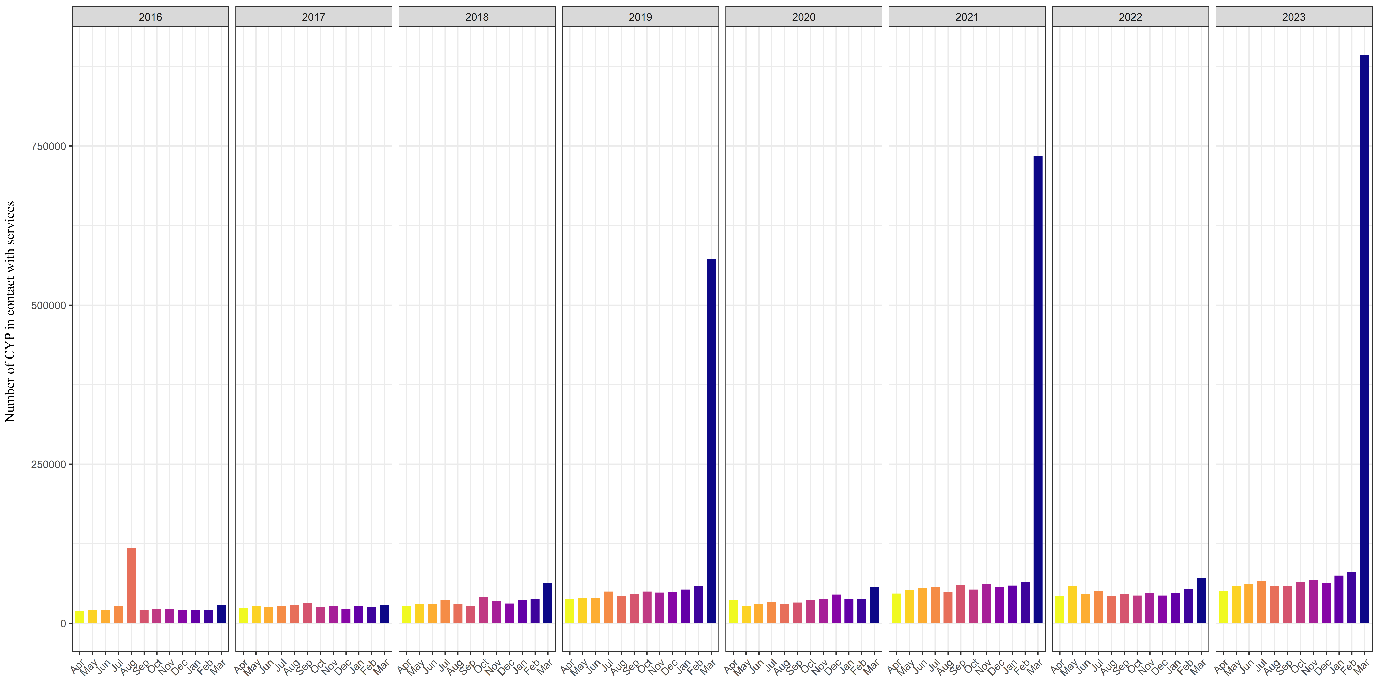

### Figure 5. Monthly number of CYP in contacts with mental health services

### Results of sensitivity analysis

In this section, we show the results related to sensitivity analyses. First, we limited our analysis to health data providers who consistently provided their records from 2016 to 2023. The spatial pattern and temporal trend of the percent of CYP in contact with mental health service and those with at least one attended care contact are reported in Figure 4 to 6.

Second, we repeated the regression model for 2021 data and found that the estimated odds ratio was generally consistent across the year. The estimated odds for 2021 data are shown in Figure 7.

.

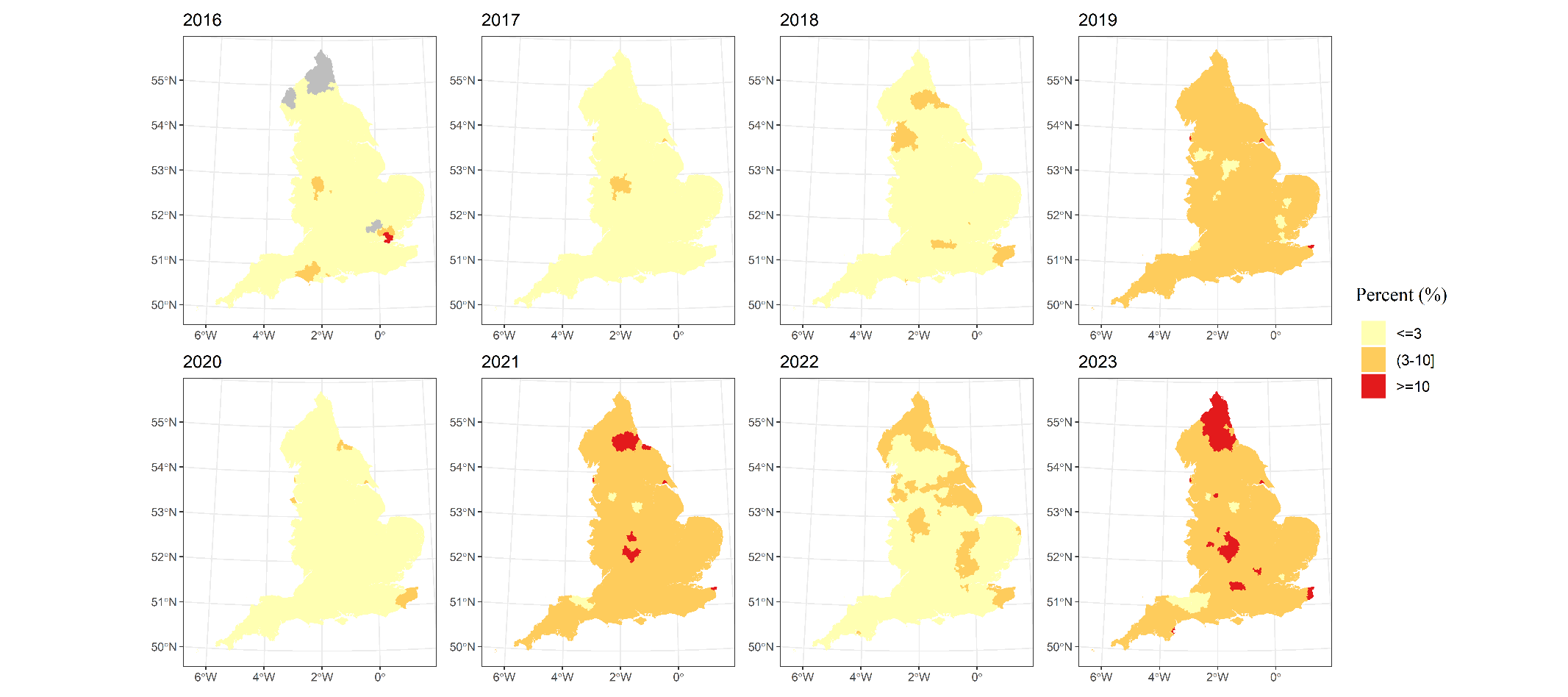

* Areas in grey indicate are local authorities where no health care provider submitted data. Additionally, three local authorities in 2016 were removed due to unreasonably high number of records.

#### Figure 5.Local authority variation in the percent children and young people in contact with mental health services in England from financial years 2016 to 2023, considering only those health providers that consistently reported over the years.

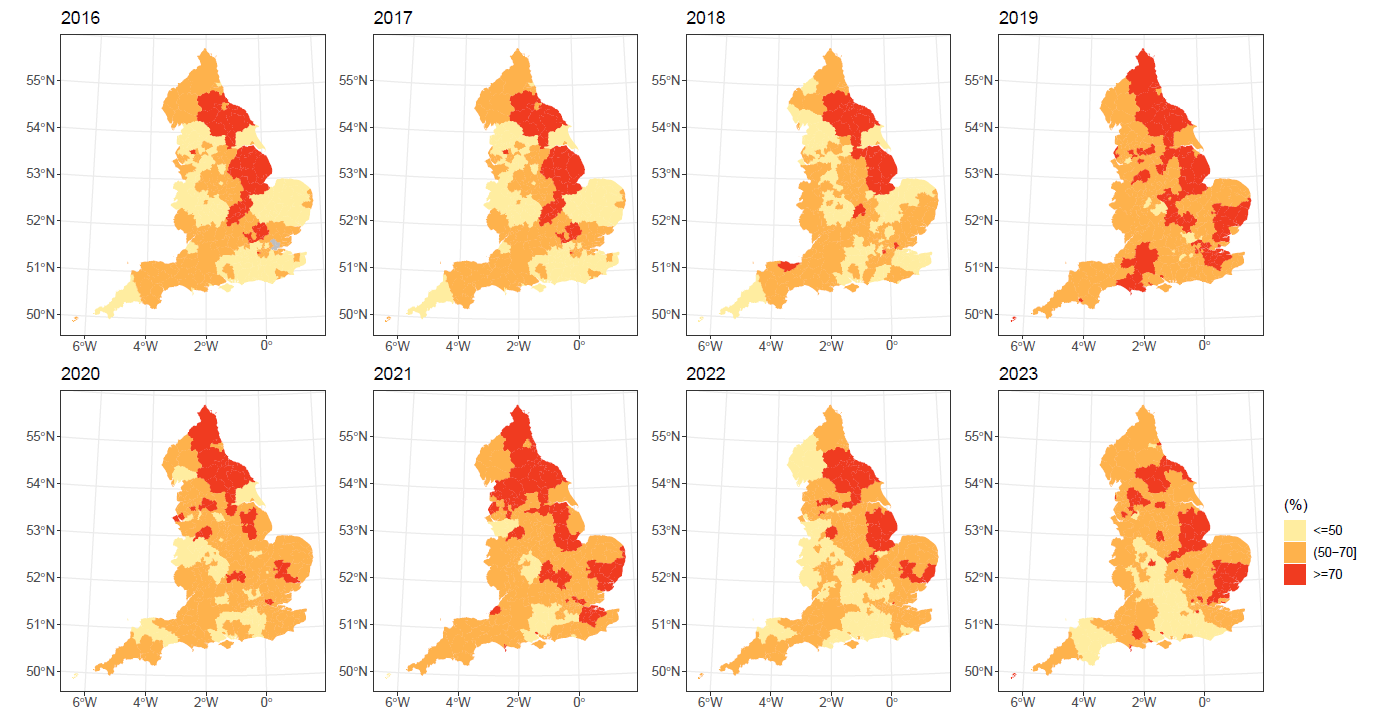

#### Figure 6.Local authority variation in the percent of children and young people with at least one attended care contact in England from financial years 2016 to 2023, considering only those local authorities that consistently reported over the years.

#### Figure 7. Temporal trend of the percent of CYP in contact with mental health service and have at least one attended care contact, considering only those local authorities that consistently reported over the years.

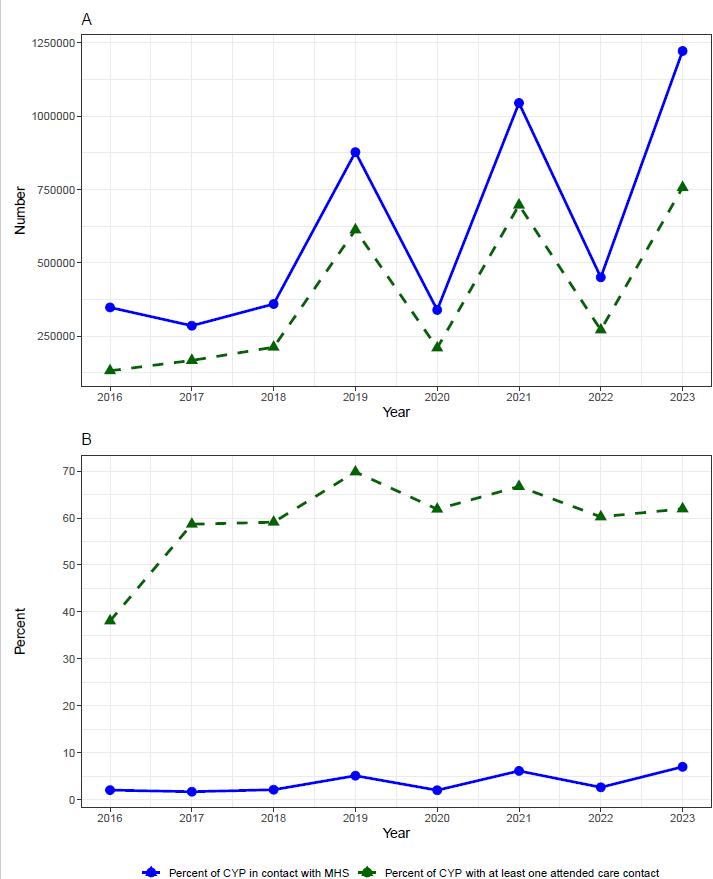
